# Increased malondialdehyde and nitric oxide formation, lowered total radical trapping capacity coupled with psychological stressors largely predict the phenome of first-episode mild depression in undergraduate students

**DOI:** 10.1101/2024.03.13.24304226

**Authors:** Francis F. Brinholi, Asara Vasupanrajit, Laura de O. Semeão, Ana Paula Michelin, Andressa K. Matsumoto, Abbas F. Almulla, Chavit Tunvirachaisakul, Decio S. Barbosa, Michael Maes

**Affiliations:** Health Sciences Graduate Program, Health Sciences Center, State University of Londrina, Londrina, Brazil; Department of Psychiatry, Faculty of Medicine, Chulalongkorn University, Bangkok, Thailand; Sichuan Provincial Center for Mental Health, Sichuan Provincial People’s Hospital, School of Medicine, University of Electronic Science and Technology of China, Chengdu, China; Key Laboratory of Psychosomatic Medicine, Chinese Academy of Medical Sciences, Chengdu, China; Medical Laboratory Technology Department, College of Medical Technology, The Islamic University, Najaf, Iraq; Cognitive Impairment and Dementia Research Unit, Faculty of Medicine, Chulalongkorn University, Bangkok, Thailand; Cognitive Fitness and Biopsychological Technology Research Unit, Faculty of Medicine Chulalongkorn University, Bangkok, Thailand; Department of Psychiatry, Medical University of Plovdiv, Plovdiv, Bulgaria; Research Institute, Medical University of Plovdiv, Plovdiv, Bulgaria; Research and Innovation Program for the Development of MU - PLOVDIV– (SRIPD-MUP), Creation of a network of research higher schools, National plan for recovery and sustainability, European Union – NextGenerationEU; Kyung Hee University, 26 Kyungheedae-ro, Dongdaemun-gu, Seoul 02447, Korea

**Keywords:** antioxidants, depression, oxidative stress, physiological stress, recurrence of illness

## Abstract

Undergraduate students are frequently afflicted by major depressive disorder (MDD). Oxidative and nitrosative stress (O&NS) has been implicated in the pathophysiology of MDD. There is no information regarding whether mild outpatient MDD (SDMD) and first episode SDMD (FE-SDMD) are accompanied by O&NS. The current study compared lipid hydroperoxides (LOOH), malondialdehyde (MDA), advanced protein oxidation products, nitric oxide metabolites (NOx), thiol groups, plasma total antioxidant potential (TRAP), and paraoxonase 1 activities among SDMD and FE-SDMD patients versus healthy controls. We found that SDMD and FE-SDMD exhibit elevated MDA and NOx, and decreased TRAP and LOOH as compared with controls. There was a significant and positive correlation between O&NS biomarkers and adverse childhood experiences (ACEs), and negative life events (NLEs). O&NS pathways, NLEs and ACEs accounted for 51.7% of the variance in the phenome of depression, and O&NS and NLS explained 42.9% of the variance in brooding. Overall, these results indicate that SDMD and FE-SDMD are characterized by reduced total antioxidant defenses and increased aldehyde and NOx production. The combined effects of oxidative and psychological stressors substantially predict the manifestation of SDMD. The differences with multi-episode MDD are attributed to specific effects of recurrence of illness and staging of illness.

## 1. Introduction

A major depressive episode (MDD) impacts a considerable portion of the global population (Browning et al., 2022). An estimated one-fifth of the global population will encounter depressive episodes during their lifetime (Malhi and Mann, 2018). Furthermore, the World Health Organization (WHO) has projected that by 2030, this condition will be the leading cause of disease burden. Mental illness is prevalent and has a significant impact on cognitive functioning, overall health, and quality of life (Kuehner, 2017).

Undergraduates at universities are frequently afflicted with this complex and pervasive disorder (Pedrelli et al., 2015). The prevalence of depression among university students is greater than that of the general population, according to numerous studies conducted in multiple countries over the past several decades (Brenneisen et al., 2016). Furthermore, the prevalence of depression in academic settings has increased over the past few decades (Ibrahim et al., 2013; Lei et al., 2016). A subsequent survey and analysis conducted in Asia revealed that between 20% and 40% of undergraduates experienced varying degrees of depression, anxiety, and tension; of these, 35% had depression levels that were higher than the average for the population (Liu et al., 2019). According to Dessauvagie et al. (2022), the median prevalence rate of depression among college students in Cambodia, Laos, Malaysia, Myanmar, Thailand, and Vietnam was 29.4%. In addition, 53% of 1455 North American college students reported experiencing depression since their freshman year, and 9% said they had contemplated suicide during that time (Furr et al., 2001). The prevalence of long-term mental health disorders among college students at three higher education institutions in the United Kingdom surpassed one-third (Stewart-Brown et al., 2000). This rate was higher than the average level reported in national surveys. Depression was reported by 21.8% of Australian college students; furthermore, their depression scores exhibited an elevation above the mean scores of the broader Australian populace (Lovell et al., 2015). The onset of depression among undergraduate university students is influenced by a multitude of risk factors. There is an involvement of personality traits as well as academic, lifestyle, social, and financial stressors (Liu et al., 2022; Mofatteh, 2020).

Oxidative and nitrosative stress (O&NS) has been implicated in the pathophysiology of MDD (Correia et al., 2023; Maes et al., 2011; 2019a; Moylan et al., 2014). O&NS is distinguished by a disparity between the antioxidant capacity of the organism and the generation of reactive oxygen and nitrosative species (ROS/RNS) (Valko et al. 2007). Multiple studies (Leonard and Maes, 2012; Maes, 1995; Maes and Carvalho, 2018a; Maes et al., 2011; 2019a) have demonstrated that MDD is distinguished by compromised lipid-associated antioxidant defenses, specifically paraoxonase 1 (PON1), heightened lipid peroxidation (as measured by lipid hydroperoxides (LOOH) and aldehyde formation (e.g., malondialdehyde (MDA), and increased production of nitric oxide metabolites (NOx) and hypernitrosylation. MDD and bipolar disorder are characterized by elevated levels of advanced protein oxidation products (AOPPs), which suggest that elevated protein oxidation may be another underlying cause of these conditions (Maes et al., 2019a).

Adverse childhood experiences (ACEs), one of the causes of MDD, is associated with elevated AOPPs, LOOH, aldehyde formation (MDA), and diminished antioxidant defenses (Moraes et al., 2018; 2019b). Psychological stressors and chronic mild stress, which are known to cause MDD and depressive-like behaviors in animal models, are characterized by induction of multiple O&NS pathways (Kubera et al., 2011; Maes et al., 2011; Moylan et al., 2014). According to the neurotoxicity theory of affective disorders (Maes et al., 2009; 2011; Moylan et al., 2013), dysfunction of gray and white matter functional plasticity, damage to astroglial and neuronal projections, and lowered neurogenesis and myelin formation result from neurotoxicity induced by activated neuro-oxidative coupled with neuro-immune pathways. In this regard, it is crucial to note that the brain is highly susceptible to oxidative damage because of its high oxygen uptake and subsequent free radical production, in addition to having a relatively weak antioxidant defense (Black et al., 2015; Camkurt et al., 2016; Gutteridge and Halliwell, 2010; Maes et al., 2011; Morris et al., 2017; Moylan et al., 2014; Palta et al., 2014). Many neurodegenerative disorders are associated with heightened O&NS, oxidative damage to proteins, DNA, and RNA, and lipid membranes (Maes et al., 2011; Popa-Wagner et al., 2013; Siwek et al., 2013).

However, the aforementioned research was conducted on patients diagnosed with MDD who had a broad range of recurrent depressive episodes, as determined by a staging index or recurrence of illness (ROI) (Maes et al., 2019b; 2023a). This is significant because a more severe phenotype of MDD is associated with increased severity of O&NS (and other pathways) and greater ROI (Maes et al., 2023a). Indeed, present evidence suggests that MDD is composed of two distinct clinical phenotypes: simple dysmood disorder (SDMD) and major dysmood disorder (MDMD). The latter phenotype is characterized by multiple interactions between biological pathway aberrations and increasing ROI (Maes et al., 2023a). However, there is currently no information regarding whether SDMD, and first episode SDMD (FE-SDMD) in particular, is accompanied by O&NS or diminished antioxidant defenses.

Hence, the objective of this research was to determine the relationship between various O&NS biomarkers and SDMD and FE-SDMD among university students as opposed to a control group of healthy individuals. In pursuit of this objective, ROS/RNS biomarkers such as LOOH, MDA, AOPP, NOx, thiol groups (-SH), plasma total antioxidant potential (TRAP), and PON1 activities were quantified. The primary hypothesis was to determine that SDMD and FE-SDMD patients have elevated O&NS levels (as indicated by increased LOOH, MDA, AOPP, and NOx) and lowered antioxidant defenses (including TRAP, PON1 activities, and -SH groups) relative to the control group.

## 2. Subjects and Methods

### 2.1. Participants

A cohort of 108 Thai-speaking individuals was enrolled in this research investigation at the outpatient Department of Psychiatry, King Chulalongkorn Memorial Hospital, Bangkok, Thailand, from November 2021 to February 2023. The participants for this research comprised individuals who met the DSM-5 criteria for MDD, as determined by senior psychiatrists. We only included students in their first episode and, thus, students with multiple episodes were excluded. Furthermore, individuals who met the inclusion criteria were those who had achieved a Hamilton Depression Rating Scale (HAM-D) score greater than 7 but less than 22, as evaluated by a clinical psychologist possessing the necessary expertise. Hence, all depressed patients included here fulfilled the diagnostic criteria of SDMD and 47 were diagnosed as FE-SDMD. This research enlisted undergraduate students from any faculty of Chulalongkorn University in Bangkok, land, between the ages of 18 and 35, of both sexes. Thus, a representative sample of undergraduate students with mild outpatient SDMD and FE-SDMD was selected.

Subjects with medical conditions such as endocrine or autoimmune disorders, psoriasis, type 1 diabetes, lupus erythematosus, chronic kidney disease, carcinoma, and psoriasis were not considered to participate. Importantly, we also excluded subjects with metabolic syndrome as diagnosed using the International Diabetes Federation and the American Heart Association/National Heart, Lung, and Blood Institute (Alberti et al., 2009). We excluded subjects who had suffered from moderate (pneumonia was present) to severe (admission to ICU) COVID-19 infection. Those who had suffered from mild COVID-19 could be included if they had not suffered from the acute infectious phase the three months prior to inclusion. Psychiatric diagnoses such as schizophrenia, substance abuse disorders involving autism spectrum disorders, alcohol or drugs (but not tobacco), anxiety and psycho-organic disorders, bipolar disorder, and schizoaffective disorder were excluded from the study. It was not permissible for expectant or lactating female undergraduate students to participate in the study. Enrolled were 62 SDMD and 47 FE-SDMD patients afflicted with an acute phase of mild depression. In this study, 44 healthy controls who had never been diagnosed with a psychiatric disorder or expressed suicidal ideation were enlisted via word-of-mouth. Controls who obtained a HAM-D score of 7 or lower were enlisted.

This investigation was reviewed and approved by the Institutional Review Board (IRB) of the Faculty of Medicine, Chulalongkorn University, Bangkok, land (IRB No.351/63). Written consent was obtained from all participants.

### 2.2. Methods

All subjects of this research responded to semi-structured interviews that inquired about sociodemographic characteristics. These characteristics included age, gender, educational attainment, current smoking status, lifetime mild COVID-19 infection history, familial psychiatric diagnoses in first-degree relatives (including major depression, bipolar disorder, anxiety, and psychosis, and suicide). The current investigation employed the following rating scales to assess the phenome of depression. The assessment of depressive symptoms and severity was conducted utilizing two instruments: the version of the HAM-D, which was translated by Lotrakul et al. (1996), and the Beck Depression Inventory-II (BDI-II), which was translated by Mungpanich (2008). The Ruminative Response Scale (RRS), developed by Thanoi et al. (2011) and translated into Thai (40) was used to assess brooding, which is a key hallmark of the phenome of depression (Vasupanrajit et al., 2023). Towards this end Vasupanrajit et al. (2023) extracted one validated principal component from the brooding items (labeled as “brooding”). The assessment of the ACEs was conducted on all participants using the ACEs Questionnaire, which was originally designed by Felitti et al. (1998). A validated translation of the ACEs Questionnaire was performed by Rungmueanporn et al. (2019). As explained by Vasupanrajit et al. (2023), composite scores (either principal component or z unit-based composites) were computed for emotional and physical abuse and neglect and sexual abuse. Utilizing the Negative Event Scale (NES), we scored the severity of total score on negative life events (NLEs) during the last year (Maybery, 2003). In the current study, we used the total sum of all items on health, finances, course interest, academic limitation, friends, boyfriend/girlfriend, parents, relatives, lecturers, and fellow students. The scale was translated into Thai and validated by Boonyamalik (2005). To calculate body mass index (BMI), measurements of height (m) and weight (kg) were obtained. DSM-5 criteria were used to diagnose tobacco use disorder (TUD).

### 2.3. Assays

Fasting blood for the analysis of O&SN biomarkers was drawn at 8:00 a.m. following an overnight fast. Aliquots of serum were kept at 80 °C until thawed for analysis. The ROS/RNS biomarkers measured include LOOH, MDA, AOPP, NOx, -SH groups, chloromethyl phenylacetatease (CMPAase), and arylesterase (AREase) activities. LOOH was measured by chemiluminescence in a Glomax Luminometer (TD 20/20), in the dark, at 30 °C for 60 min (Flecha et al., 1991; Panis, et al., 2012), and the findings were represented in relative light units. Complexation with two molecules of thiobarbituric acid and MDA estimation through high-performance liquid chromatography (HPLC) (HPLC Alliance e2695, Waters’, Barueri, SP, Brazil) were used to assess MDA levels (Bastos et al., 2012) (Column Eclipse XDB-C18, Agilent, Santa Clara, CA, USA; mobile phase composed of 65% potassium phosphate buffer (50 mM pH 7.0) and 35% HPLC grade methanol; flow rate of 1.0 mL/min; temperature of 30 degrees Celsius; wavelength of 532 nm). Based on a calibration curve, the MDA concentration in the samples was determined and is given as mmol of MDA per mg of protein. At 340 nm, AOPP was quantified in mM of equivalent chloramine T using a microplate reader (EnSpire, Perkin Elmer, Waltham, MA, USA) (Hanasand et al., 2012; Witko-Sarsat et al., 1996). NO metabolites (NOx) were determined by measuring nitrite and nitrate concentrations in a microplate reader (EnSpire®, Perkin Elmer, USA) at 540 nm (Navarro-Gonzálvez et al., 1998), and the results are reported as M. The -SH groups were evaluated using a microplate reader (EnSpire®, Perkin Elmer, USA) at 412 nm, and the results were expressed in millimolar (M) (Hu, 1994; Taylan and Resmi, 2010). All inter-assay coefficients of variation were <10.0%). PON1 status was determined by AREase and CMPAase activity and PON1 Q192R genotypes in the present investigation (Brinholi et al., 2023; Maes et al., 2022; Matsumoto et al., 2021; Moreira et al., 2017; 2019). We investigated the rate of phenylacetate hydrolysis at low salt concentration by measuring the activity of AREase and CMPAase (Sigma, St. Louis, MO, USA) ((Brinholi et al., 2023; Maes et al., 2022; Matsumoto et al., 2021; Moreira et al., 2017; 2019). A Perkin Elmer® EnSpire model microplate reader (Waltham, MA, USA) was used to monitor the rate of phenylacetate hydrolysis at a constant temperature of 25 °C for 4 min (16 measurements with 15 s between each reading). The activity was expressed in units per milliliter (U/mL) based on the phenyl acetate molar extinction coefficient of 1.31 mMol/L cm^−1^. CMPA and phenyl acetate were also utilized to stratify the functional genotypes of the PON1Q192R polymorphism (PON1 192Q/Q, PON1 192Q/R, and PON1 192R/R) (Sigma, PA, USA). The phenylacetate reaction is conducted with high salt concentrations, which partially inhibits the R allozyme’s activity, allowing for a sharper distinction between the three functional genotypes. TRAP was measured by chemiluminescence as described by Repetto et al. (1996). The 2,2’ azo-bis generates peroxyl radicals rapidly via interaction with carbon-centered radicals and molecular oxygen. These free radicals react with luminol (which acts as an amplifier), producing chemiluminescence. The addition of plasma reduces the chemiluminescence at baseline levels for a period (induction time) proportional to the concentration of TRAP until free radicals are regenerated, returning to initial levels of chemiluminescence. This experiment was conducted in a Beckman β counter, model LS6000 (Fullerton, CA, USA) in anon-coincident-counting mode for 25 minutes and with a response between 300 and 620 nm.

### 2.4. Statistical analyses

The statistical analysis for this investigation was performed using version 29 of IBM SPSS for Windows. The X^2^-test (or Fisher’s exact probability test) was employed by the researchers as a contingency table analysis to assess the statistical relationships between categorical variables. Pearson’s product-moment correlation coefficients were employed to determine the statistical relationships between continuous variables. The researchers utilized analysis of variance to examine the associations between diagnostic categories and clinical data. Multivariate GLM analysis was employed to decipher the associations between the O&NS biomarkers and the diagnosis of (FE-)SDMD versus controls. Effects were adjusted for confounding variables when significant by entering these variables as covariates. To determine the effect of O&NS biomarkers coupled with ACEs, NLEs, and demographic characteristics on the clinical features, manual multiple regression analysis was applied. In addition, an automatic forward stepwise regression technique was implemented. A significance level of p=0.05 was set for the inclusion of variables, while p=0.07 was determined for their exclusion. The model statistics comprised F statistics and their respective p-values, and the effect size was assessed using partial Eta squared or R^2^. Standardized β coefficients, t-statistics (with p-values) were computed for every variable that was included in the final regression models. To ascertain the existence of heteroskedasticity, the White test and a modified variant of the Breusch-Pagan test were executed. We examined the data for possible multicollinearity and collinearity using a tolerance limit of 0.25 and a variance inflation factor threshold of four or greater. In addition, for this analysis, we employed the condition index and variance proportions that were extracted from the table of collinearity diagnostics. Each of the analyses was performed utilizing two-tailed tests. We considered a p-value (alpha level) of 0.05 or less to be indicative of statistical significance. Where necessary, we utilized transformations such as logarithmic, square-root, and rank-based inversed normal to ensure that our data indicators followed a normal distribution. Binary logistic regression analysis was performed using the SDMD diagnosis as dependent variable (controls as reference group). We computed the Odds ratio with 95% confidence intervals, the accuracy and Nagelkerke metrics. The latter was used as effect size. The O&NS biomarkers and demographic data were used as explanatory variables. We used a stepwise automatic and manual method to delineate the best predictors. We used a p-to-enter of 0.05, and a p-to-remove of 0.07. Multiple predictors were combined into one variable using z unit-based composite scores. The primary statistical analysis was the multiple regression analysis with the SDMD phenome as dependent variable and O&NS biomarkers, ACEs and NLEs as explanatory variables. A priori, it was assumed that an explained variance of around 15% would be relevant. G*Power 3.1.9.4 was applied to ascertain the sample size a priori. Using an effect size of f = 0.176, a power of alpha = 0.05, power of 0.8 and a maximum number of predictors of 6, the a priori estimated sample size was 84.

## 3. Results

### 3.1. Sociodemographic and clinical data

**Table 1** shows the socio-demographic and clinical data of the SDMD patients and controls. There were no significant differences between both groups in age, education, body mass index (BMI), sex ratio, relationship status, TUD, student alcohol drinking, and mild COVID-19 history. Patients with SDMD showed significantly increased ACE scores of emotional abuse, physical abuse, sexual abuse, emotional neglect, and physical neglect, negative life events, HAM-D, BDI-II, brooding, and phenome scores as compared with controls.

**Table 1.** – Clinical data of healthy controls (HC) and simple dysmood disorder (SDMD) patients.

| Variables | HC<br>(n=44) | SDMD<br>(n=64) | F/X<sup>2</sup> | df | p |
| --- | --- | --- | --- | --- | --- |
| Age (years) | 23.48 ± 3.18 | 22.45 ± 3.15 | 2.75 | 1/106 | 0.100 |
| Education (years) | 16.57 ± 1.49 | 15.92 ± 2.65 | 2.15 | 1/106 | 0.146 |
| BMI (k/m<sup>2</sup>) | 22.33 ± 3.57 | 22.41 ± 5.13 | 0.01 | 1/106 | 0.927 |
| Sex (Male/Female) | 37 / 07 | 54 / 10 | 0.01 | 1 | 0.968 |
| Status (Single/relationship) | 27 / 17 | 27 / 37 | 3.84 | 1 | 0.050 |
| TUD (no/yes) | 43 / 1 | 59 / 5 | FFHET | - | 0.398 |
| Student alcohol drinking (no/yes) | 41 / 3 | 53 / 11 | 2.48 | 1 | 0.115 |
| Mild Covid-19 history (no/yes) | 30 / 14 | 49 / 15 | 0.93 | 1 | 0.334 |
| ACE sum score Emotional Abuse | 0.07 ± 0.33 | 0.53 ± 0.73 | KWT | - | <0.001 |
| ACE sum score Physical Abuse | 0.16 ± 0.48 | 0.41 ± 0.68 | KWT | - | 0.041 |
| ACE sum score Sexual Abuse | 0.30 ± 0.93 | 0.75 ± 1.27 | KWT | - | 0.045 |
| ACE sum score Emotional Neglect | 9.39 ± 4.50 | 13.73 ± 4.86 | KWT | - | <0.001 |
| ACE sum score Physical Neglect | 7.41 ± 2.27 | 8.53 ± 2.67 | KWT | - | 0.025 |
| Negative Life Events | 42.20 ± 28.11 | 76.64 ± 27.27 | KWT | - | <0.001 |
| HAM-D | 2.39 ± 1.96 | 14.64 ± 4.79 | KWT | - | <0.001 |
| BDI-II | 6.77 ± 6.49 | 24.67 ± 11.54 | KWT | - | <0.001 |
| Brooding (z scores) | -0.89 ± 0.80 | 0.44 ± 0.69 | KWT | - | <0.001 |
| Phenome (z scores) | -2.62 ± 1.28 | 1.80 ± 1.85 | KWT | - | <0.001 |
| PON1 genotypes: QQ / QR / RR | 8 / 16 / 20 | 8 / 30 / 24 | 1.61 | 2 | 0.446 |

Values are presented as mean ± SD or as a ratio; F: results of analysis of variance; $\chi^2$ : results of contingency analysis; KWT: results of the Kruskal-Wallis one-way analysis of variance; FFHET: Fisher-Freeman-Halton Exact Test. BMI: body mass index; TUD: tobacco use disorder; ACE: adverse childhood experiences; HAM-D: Hamilton Depression Rating Scale; BDI-II: Beck's Depression Inventory-II.

### 3.2. O&NS biomarkers in SDMD, FE-SDSMD and controls

**Table 2** shows the results of multivariate GLM analysis that examines the associations between of O&NS biomarkers and the diagnosis SDMD after covarying for age and sex (BMI was not significant). The same table also shows the model-generated estimated marginal mean values of the O&NS biomarkers. We found that after covarying for the significant effects of age and sex, there was a significant association between SDMD and the O&NS biomarkers. Analyses of parameter estimates show that TRAP and LOOH were significantly lower in patients than in controls, whereas MDA and NOx were significantly increased in SDMD. All significances shown in Table 2 remained significant at p=0.05 after False Discovery Rate (FDR) p correction, except NOx (p=0.053). Table 2 and Table 1 (see PON1 genotypes) show that there were no significant alterations in total PON1 status (enzyme activities and PON1 genotype distribution) between both groups. **Electronic Supplementary File (ESF) 1** shows that the same findings were observed in FE-SDMD. False discovery rate p correction showed that all significant differences between FE-SDMD and controls remained significant after p correction, including NOx (p=0.0432) and MDA (p=0.0337).

**Table 2.**
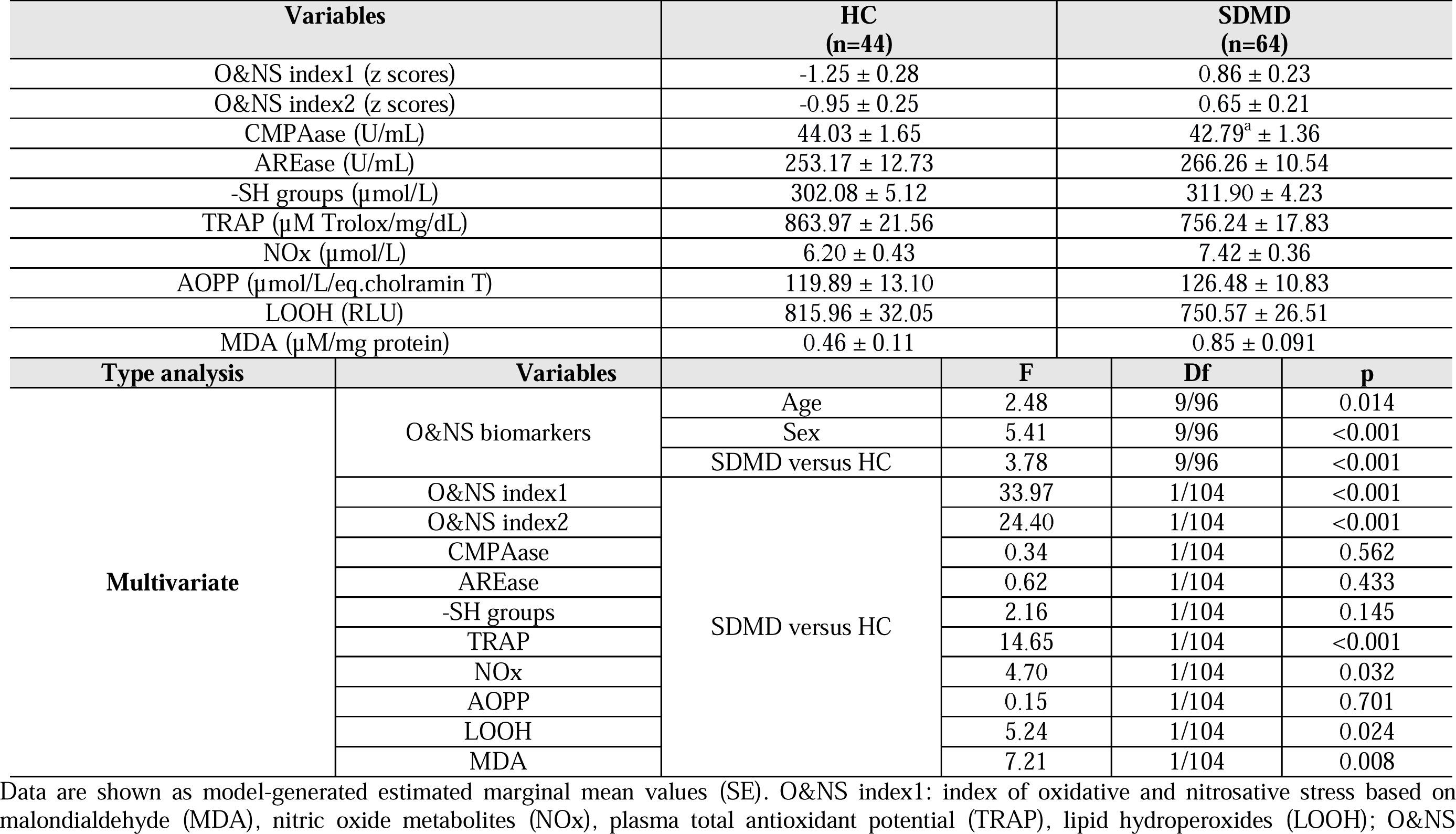
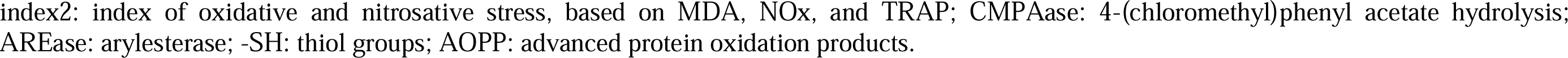
Results of general linear models (GLM) which show the associations between oxidative and nitrosative stress biomarkers and simple dysmood disorder (SDMD) versus healthy controls (HC). The associations were adjusted for age and sex.

| Variables |  | HC<br>(n=44) | SDMD<br>(n=64) |  |  |
| --- | --- | --- | --- | --- | --- |
| O&NS index1 (z scores) |  | -1.25 ± 0.28 | 0.86 ± 0.23 |  |  |
| O&NS index2 (z scores) |  | -0.95 ± 0.25 | 0.65 ± 0.21 |  |  |
| CMPAase (U/mL) |  | 44.03 ± 1.65 | 42.79<sup>a</sup> ± 1.36 |  |  |
| AREase (U/mL) |  | 253.17 ± 12.73 | 266.26 ± 10.54 |  |  |
| -SH groups (μmol/L) |  | 302.08 ± 5.12 | 311.90 ± 4.23 |  |  |
| TRAP (μM Trolox/mg/dL) |  | 863.97 ± 21.56 | 756.24 ± 17.83 |  |  |
| NOx (μmol/L) |  | 6.20 ± 0.43 | 7.42 ± 0.36 |  |  |
| AOPP (μmol/L/eq.cholramin T) |  | 119.89 ± 13.10 | 126.48 ± 10.83 |  |  |
| LOOH (RLU) |  | 815.96 ± 32.05 | 750.57 ± 26.51 |  |  |
| MDA (μM/mg protein) |  | 0.46 ± 0.11 | 0.85 ± 0.091 |  |  |
| Type analysis | Variables |  | F | Df | p |
| Multivariate | O&NS biomarkers | Age | 2.48 | 9/96 | 0.014 |
|  |  | Sex | 5.41 | 9/96 | <0.001 |
|  |  | SDMD versus HC | 3.78 | 9/96 | <0.001 |
|  | O&NS index1 | SDMD versus HC | 33.97 | 1/104 | <0.001 |
|  | O&NS index2 |  | 24.40 | 1/104 | <0.001 |
|  | CMPAase |  | 0.34 | 1/104 | 0.562 |
|  | AREase |  | 0.62 | 1/104 | 0.433 |
|  | -SH groups |  | 2.16 | 1/104 | 0.145 |
|  | TRAP |  | 14.65 | 1/104 | <0.001 |
|  | NOx |  | 4.70 | 1/104 | 0.032 |
|  | AOPP |  | 0.15 | 1/104 | 0.701 |
|  | LOOH |  | 5.24 | 1/104 | 0.024 |
|  | MDA |  | 7.21 | 1/104 | 0.008 |

Data are shown as model-generated estimated marginal mean values (SE). O&NS index1: index of oxidative and nitrosative stress based on malondialdehyde (MDA), nitric oxide metabolites (NOx), plasma total antioxidant potential (TRAP), lipid hydroperoxides (LOOH); O&NS
index2: index of oxidative and nitrosative stress, based on MDA, NOx, and TRAP; CMPAase: 4-(chloromethyl)phenyl acetate hydrolysis; AREase: arylesterase; -SH: thiol groups; AOPP: advanced protein oxidation products.

There were no significant effects of the drug state of the patients on any of the O&NS variables, even without FDR p correction, namely sertraline (n=20, F=1.54, df=10/92, p=0.138), fluoxetine (n=10, F=1.00, df=10/92, p=0.443), escitalopram (n=14, F=0.569, df=10/92, p=0.835), benzodiazepines (n=27, F=0.542, df=10/92, p=0.856), and antipsychotics (n=12, F=1.24, df=10/92, p=0.279).

### 3.3. Prediction of SDMD and FE-SDMD using O&NS biomarkers

**Table 3** shows the results of binary logistic regression analyses with the diagnosis as dependent variable and the biomarkers as explanatory variables. First, we entered all biomarkers in the regression analysis. Model #1 shows that SDMD was predicted by a combination of MDA and NOx (both positively) and LOOH, age and TRAP (all three inversely) (Χ^2^=31.21, df=5, p<0.001; accuracy=71.4% and Nagelkerke value of 0.387). Based on these results we have constructed two post-hoc O&NS composite scores as a feature reduction method because both logistic regression and multiple regression analyses showed that MDA and NOx were positively associated with depression (either diagnosis or severity), whereas in the same regression equation LOOH and TRAP were inversely associated. The first composite is: z score of MDA (z MDA) + z NOx – z TRAP – z LOOH (O&NS index1). As described in our discussion, lowered LOOH values may be clinically relevant. Nevertheless, we constructed a second composite without LOOH as: z MDA + z NOx – zTRAP (O&NS index2). Table 2 shows that both indices were significantly higher in SDMD than in controls. Model #2 (Χ^2^=30.15, df=2, p<0.001; accuracy=72.5% and Nagelkerke of 0.376) and model #3 (Χ^2^=23.82, df=2, p<0.001; accuracy=70.3% and Nagelkerke of 0.307) show that using both indices instead of the 4 biomarkers, the results (effect size) did not change. Model #4 shows that MDA (positively) combined with LOOH, and age (both inversely) significantly predicted SDMD (Χ^2^=23.38, df=3, p<0.001; accuracy=65.9% and Nagelkerke of 0.302). Introducing ACEs and NLEs in the equation (model #5) shows that NLEs, ACEs and the O&NS index1 highly significantly predict the diagnosis (Χ^2^=53.12, df=3, p<0.001; accuracy=79.1% and Nagelkerke of 0.590). Model #6 shows that NOx and MDA (positively), age and LOOH (inversely) predicted FE-SDMD (Χ^2^=27.99, df=4, p<0.001; accuracy=67.8% and Nagelkerke of 0.357).

**Table 3.** Results of logistic regression analyses with the diagnoses of simple dysmood disorder (SDMD) and first episode SDMD (FE-SDMD) in outpatients as dependent variable (normal controls as reference group)

| Models | Explanatory variables | B | SE | W (df=1) | p | OR | 95% confidence intervals |  |
| --- | --- | --- | --- | --- | --- | --- | --- | --- |
|  |  |  |  |  |  |  | Lower | higher |
| **Model #1** | **NOx** | 0.546 | 0.275 | 3.95 | 0.047 | 1.720 | 1.008 | 2.958 |
|  | **TRAP** | -0.498 | 0.269 | 3.43 | 0.064 | 0.613 | 0.359 | 1.030 |
|  | **MDA** | 0.938 | 0.368 | 6.50 | 0.011 | 2.552 | 1.242 | 5.252 |
|  | **LOOH** | -0.753 | 0.297 | 6.45 | 0.011 | 0.471 | 0.263 | 0.842 |
|  | **Age** | -0.234 | 0.094 | 6.27 | 0.012 | 0.797 | 0.657 | 0.950 |
| **Model #2** | **Age** | -0.239 | 0.092 | 6.69 | 0.010 | 0.788 | 0.657 | 0.944 |
|  | **O&NS index1** | 1.326 | 0.323 | 16.81 | <0.001 | 3.765 | 1.998 | 7.096 |
| **Model #3** | **Age** | -0.220 | 0.086 | 6.56 | 0.010 | 0.803 | 0.678 | 0.950 |
|  | **O&NS index2** | 1.122 | 0.298 | 14.14 | <0.001 | 3.072 | 1.711 | 5.515 |
| **Model #4** | **Age** | -0.182 | 0.083 | 4.81 | 0.028 | 0.834 | 0.708 | 0.981 |
|  | **MDA** | 1.027 | 0.363 | 8.01 | 0.005 | 2.792 | 1.371 | 5.685 |
|  | **LOOH** | -0.847 | 0.274 | 9.57 | 0.002 | 0.429 | 0.251 | 0.733 |
| **Model #5** | **ACEs** | 0.674 | 0.348 | 3.76 | 0.053 | 1.963 | 0.992 | 3.882 |
|  | **NLEs** | 0.044 | 0.012 | 12.91 | <0.001 | 1.045 | 1.020 | 1.071 |
|  | **O&NS index1** | 1.378 | 0.375 | 13.51 | <0.001 | 3.966 | 1.903 | 8.269 |
| **Model #6** | **Age** | -0.237 | 0.094 | 6.04 | 0.011 | 0.789 | 0.657 | 0.948 |
|  | **NOx** | 0.534 | 0.269 | 3.93 | 0.047 | 1.706 | 1.006 | 2.892 |
|  | **MDA** | 0.976 | 0.367 | 7.06 | 0.008 | 2.653 | 1.292 | 5.448 |
|  | **LOOH** | -0.907 | 0.284 | 10.22 | 0.001 | 0.404 | 0.231 | 0.704 |

O&NS index1: index of oxidative and nitrosative stress based on malondialdehyde (MDA), nitric oxide metabolites (NOx), plasma total antioxidant potential (TRAP), lipid hydroperoxides (LOOH); O&NS index2: index of oxidative and nitrosative stress, based on MDA, NOx, and TRAP; ACE: adverse childhood experiences; NLE: negative life events. Models #1 to 5: SDMD versus controls; Model #6: FE-SDMD versus controls.

### 3.4. Associations between the O&NS indices and clinical data

**Table 4** shows the intercorrelation matrix between the O&NS index1 and index2 and phenome data. Both baseline O&NS index1 and O&NS index2 were significantly and positively correlated with emotional neglect, sexual abuse, HAM-D, BDI-II, brooding, and the phenome, but not with emotional and physical abuse, and physical neglect. Negative life events were significantly and positively correlated with the O&NS index1. MDA was positively associated with all 4 clinical phenome data and with emotional neglect. LOOH was inversely associated with negative life events, HAM, BDI-II and the phenome scores.

**Table 4.** Intercorrelation matrix between oxidative and nitrosative stress (O&NS) biomarkers and clinical variables.

| Variables | O&NS index1 | O&NS index2 | MDA | LOOH |
| --- | --- | --- | --- | --- |
| Emotional Neglect | 0.252(0.009) | 0.201(0.037) | 0.231(0.016) | -0.129(0.183) |
| Sexual Abuse | 0.239(0.013) | 0.245(0.011) | 0.101(0.300) | -0.022(0.820) |
| Negative Life Events | 0.240(0.012) | 0.123(0.206) | 0.081(0.406) | -0.244(0.011) |
| HAM-D | 0.358(<0.001) | 0.252(0.008) | 0.257(0.007) | -0.257(0.007) |
| BDI-II | 0.328(<0.001) | 0.253(0.008) | 0.235(0.014) | -0.203(0.035) |
| Brooding | 0.368(<0.001) | 0.286(0.003) | 0.211(0.029) | -0.170(0.079) |
| Phenome | 0.368(<0.001) | 0.277(0.004) | 0.261(0.006) | -0.241(0.012) |

O&NS index1: index of oxidative and nitrosative stress based on malondialdehyde (MDA), nitric oxide metabolites (NOx), plasma total antioxidant potential (TRAP), lipid hydroperoxides (LOOH); O&NS index2: index of oxidative and nitrosative stress, based on MDA, NOx, and TRAP; HAM-D: Hamilton Depression Rating Scale; BDI-II: Beck's Depression Inventory-II. These associations are computed in controls and SDMD patients.

### 3.5. Results of multiple regression analyses

**Table 5** displays the results of various regression models (performed on SDMD patients) using the phenome, brooding, total BDI-II, and total HAM-D as dependent variables. When entering the O&NS data and demographic data (Table 4, regression #1a), the phenome was best predicted by O&NS index1 (positively) and age (inversely). **Figure 1** shows the partial regression of the phenome score on the O&NS index1 after considering the effects of age. In subjects with FE-SDMD we found that the same input variables explained 21.4% of the variance in the phenome of FE-DSMD (F=11.81, df=2/87, p<0.001). Table 5, regression #1b shows that after considering the effects of NLEs and total ACE score, the effects of the O&NS index1 remained significant, and that all three variables together explained 51.7% of the variance in the phenome. Regression #1c excluded patients with more than one episode and detected that 55.7% of the variance in the FE-SDMD phenome was explained by NLEs and O&NS index1. Model #2a shows that 13.5% of the variance in brooding was explained by the O&NS index1. Model #2b shows that 42.9% of the variance was explained by this index and NLEs (the ACE score was not significant). Model #3 shows that 42.1% of the variance in the BDI-II score was explained by NLEs and MDA (both positively). Model #4 displays that 38.0% of the variance in the total HAM-A score was explained by NLEs, MDA (both positively) and LOOH (inversely).

**Table 5.**
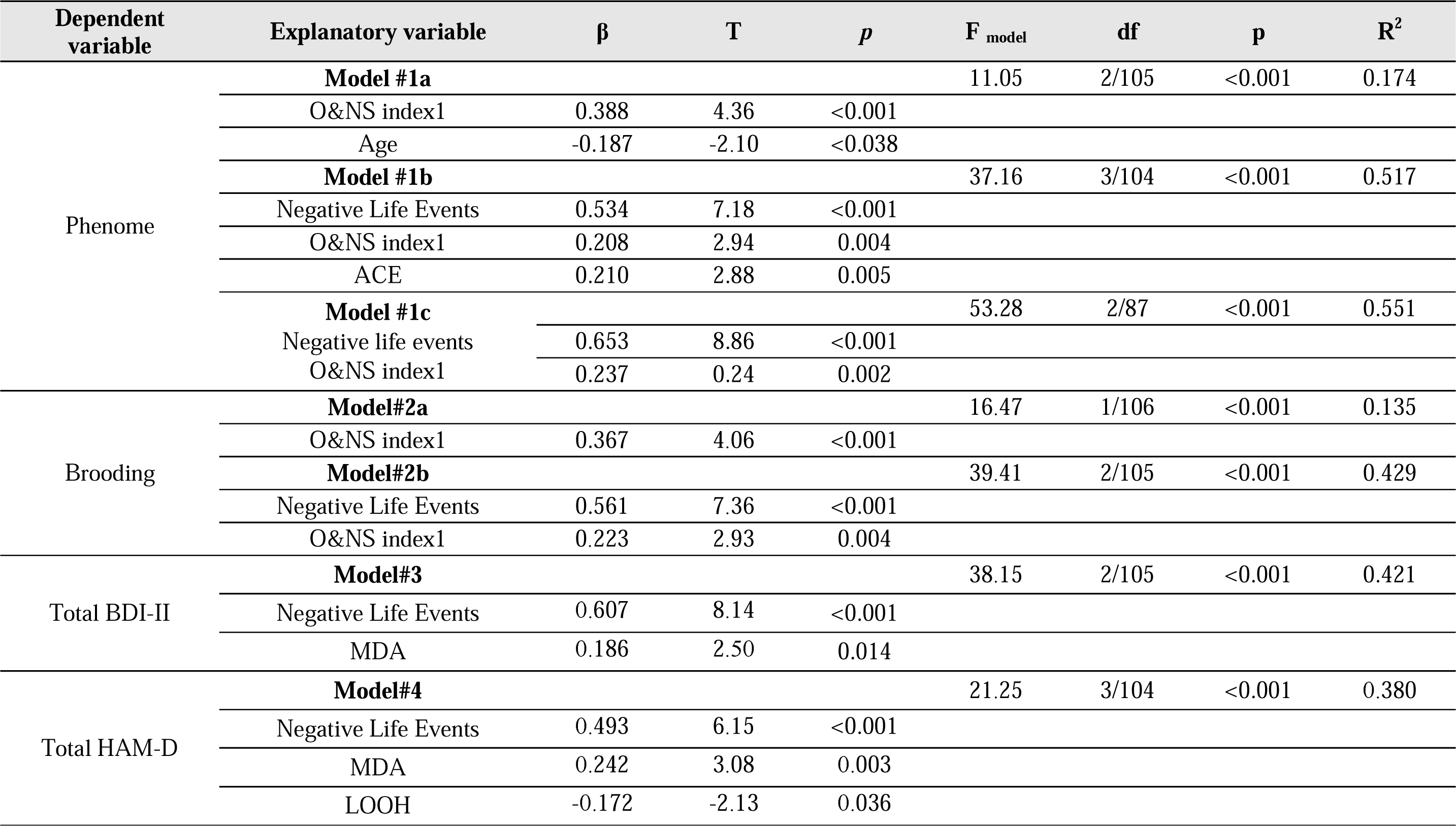
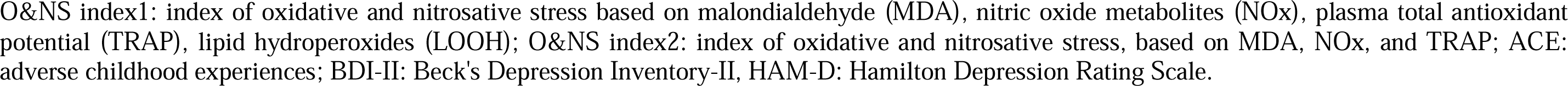
Results of multiple regression analysis with phenome data as dependent variables and oxidative and nitrosative stress (O&NS) biomarkers, adverse childhood experiences, and negative life events as explanatory variables.

**Figure 1.**
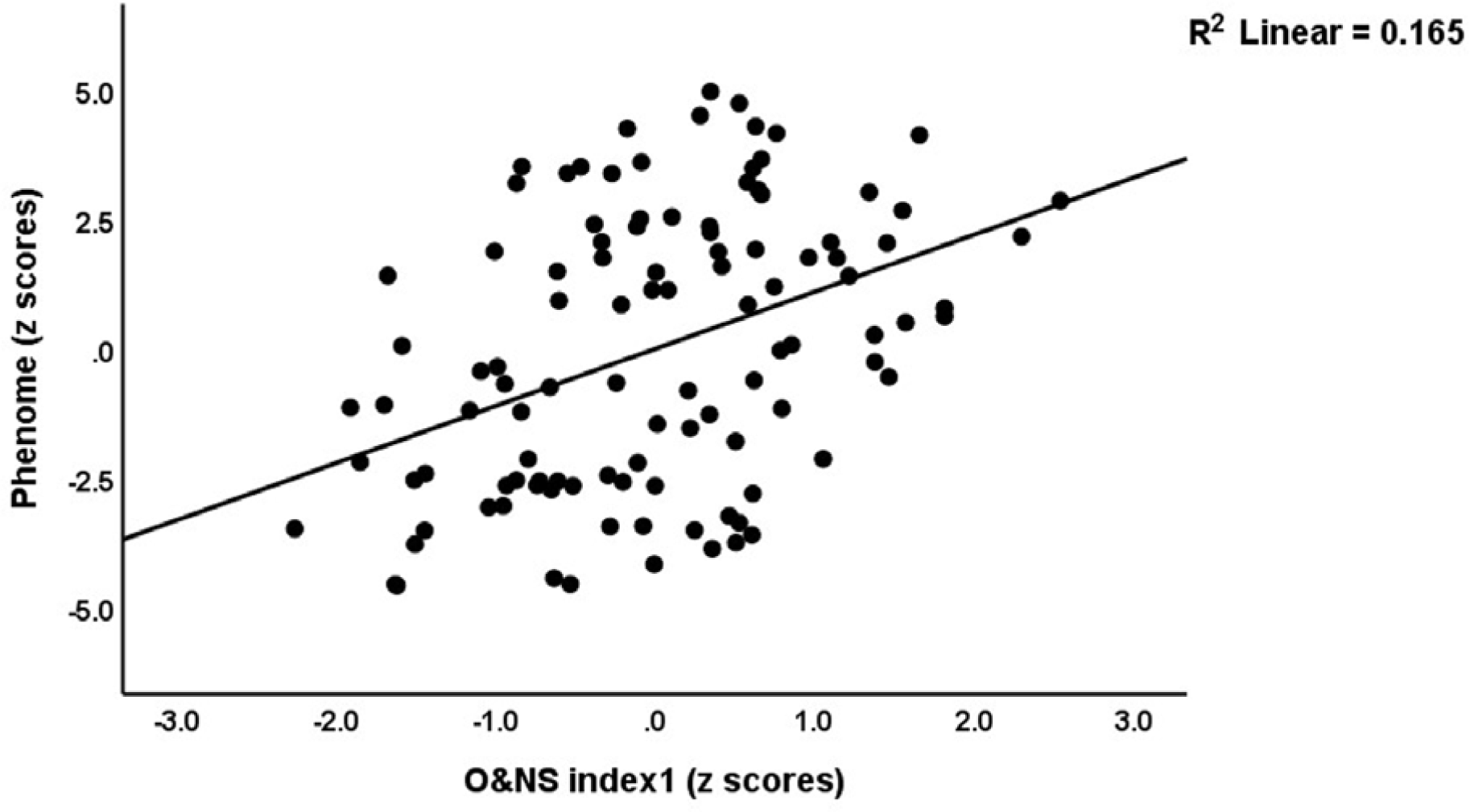
Partial regression of the phenome score on an index of oxidative and nitrosative stress (O&NS index1) after considering the effects of age.

## 4. Discussion

### 4.1. Changes in O&NS in SDMD and FE-SDMD

The primary discovery of this research is that SDMD and FE-SDMD exhibits indicators of elevated O&NS, such as elevated levels of MDA and NOx, and decreased levels of TRAP. Overall, these results indicate that (FE-)SDMD is characterized by reduced total antioxidant defenses and increased aldehyde and NO production.

Increased O&NS is a key pathophysiologic factor in MDD, particularly in recurrent MDD and MDMD, according to previous research (Maes et al., 2011; 2019b; 2023a; Moylan et al., 2014). As an illustration, research has found that patients who have been diagnosed with recurrent MDD exhibit heightened concentrations of MDA (Rybka et al., 2013). Furthermore, depressed individuals who have experienced recurrent episodes of depression demonstrate even higher levels of MDA concentrations than those experiencing their initial episodes (Stefanescu and Ciobica, 2012). Clinical research has demonstrated elevated levels of NOx (Gomes et al., 2018; Maes et al., 2019a) and NO accumulation, as evidenced by increased protein nitrosylation (Maes et al., 2008; 2013). Furthermore, chronic MDD is associated with an additional increase in nitrosylation patterns (Maes et al., 2013). Moreover, the extent of nitrosylation remains constant while antidepressant treatment is ongoing, whereas there is evidence suggesting that treatment mitigates the detrimental effects of oxidative stress (Maes and Leunis, 2014). The latter findings indicate that staging variables, such as chronicity of depression and treatment resistance, might be associated with increased nitrosylation and NO production. As previously discussed, a substantial body of literature indicates that MDD is associated with diminished antioxidant defenses and total antioxidant capacity (including TRAP) ((Maes et al., 2011; Moylan et al., 2014) and ROI is significantly correlated with diminished activity of particular antioxidant levels (Maes et al., 2018b).

All in all, based on the results obtained from this study, SDMD and FE-SDMD exhibit similarities to multiple episode MDD and MDMD, including increased MDA and NOx production, and lowered TRAP levels. Furthermore, none of the students included here showed metabolic syndrome. This is important as metabolic syndrome is (independently from mood disorders) characterized by increased MDA and AOPP (Morelli et al., 2021). Therefore, increased MDA and NOx formation as well as lowered TRAP in SDMD occur independently from metabolic syndrome.

### 4.2. Findings not in accordance with the a priori hypothesis

In contrast to the a priori hypothesis, SDMD and FE-SDMD are not accompanied by increased AOPP and LOOH levels, and by lowered -SH groups and activities of the two catalytic sites of the PON1 enzyme (Brinholi et al. 2023; Maes et al., 2011; Moreira et al., 2019). Another unexpected finding is that both SDMD and FE-SDMD were associated with decreased concentrations of LOOH.

An evaluation of the catalytic activity of PON1 can be conducted using a range of substrates, including AREase activity (phenylacetate or 4(p)-nitrophenyl acetate) and paraoxonase/phosphotriesterase activity (with CMPA or paraoxon as substrates) (Ceron et al., 2014). Moreira et al. (2019) reported that, in mood disorders, ROI as measured by the number of previous depressive and manic episodes, is significantly correlated with decreased total PON1 and CMPAase activities. Therefore, it is difficult to draw comparisons between the findings of this study and those of the aforementioned research due to substantial differences in the study populations: Brinholi et al. (2023) and Moreira et al. (2019) examined recurrent MDD and bipolar depression, while we examined (FE-)SDMD. In addition, it is difficult to compare our findings with most prior publications on PON1 activities because most of those publications did not include both enzyme activities of PON1 or drew incorrect conclusions regarding its catalytic sites (Moreira et al., 2019). It can be inferred that reduced PON1 activities accurately indicate increasing ROI explaining that no changes may be found in SDMD. Hence, we may posit that the PON1 system including the PON1 Q198R gene variant is not really associated with depression as such, but rather with one of its features, namely ROI. The differences in PON1 activities between the current study and previous research in MDD may, therefore, be explained by differences in the study samples that are associated with ROI, namely FE-SDMD versus multi-episode MDD in previous studies.

Plasma -SH concentrations in (FE-)SDMD patients were not significantly reduced, contrary to expectations, despite TRAP inhibition. Previous studies have identified decreased thiol groups in patients with MDD or depressive symptoms (Beğinoğlu et al., 2023; Roomruangwong et al., 2017; Tuncay et al., 2023). Conversely, several studies have detected elevations in -SH groups in patients with depression and cognitive impairment, suggesting that increased O&NS levels resulted in increased plasma thiols (Gałecki et al., 2013; Karaaslan et al., 2018; Sundaram and Prabhu, 2021). The consequence is a lack of consensus concerning the status of -SH groups in MDD. This may be explained, as a multitude of confounding factors, including but not limited to reproductive hormones, nutrition, body mass index, smoking status, and sleep regimen, have the potential to exert an influence on thiol groups (Baykan et al., 2018; Moylan et al., 2013; Roomruangwong et al., 2020).

In addition to MDA and other aldehydes, elevated concentrations of AOPP (Gomes et al., 2018) and LOOH (Maes et al., 2019a) have been identified in some studies on MDD (Maes et al., 2011). An increase in AOPP concentrations is a biomarker of chlorinative stress and protein oxidation (Moylan et al., 2014). Consequently, FE-SDMD may manifest initially as increased lipid peroxidation accompanied by the formation of aldehydes, while protein oxidation may be limited to more severe or recurring depression.

At first glance, the disparity between the substantially elevated levels of secondary lipid peroxidation markers (MDA) and the diminished levels of primary lipid peroxidation products (LOOH) may seem to contradict one another. Extracellular fluids consist of lipoproteins and membranes, both of which contain lipophilic antioxidants that are susceptible to chemical reactions with peroxyl lipid radicals (Catalá, 2006). Antioxidants, when present in low concentrations, can inhibit or significantly delay the oxidation of an oxidizable substrate (Haliwell, 1990; 1995). Consequently, a reduction in LOOH detection may occur because of the chain reaction being terminated when the interaction between a lipid radical and an antioxidant molecule obstructs the emission of light (Catalá, 2006). As a result, the interpretation of LOOH quantitation may sometimes pose difficulties, whereas the measurement of MDA enables a more accurate estimation of lipid peroxidation (Morales and Munne-Bosch, 2019). By utilizing reverse phase HPLC techniques, MDA may be differentiated from other interfering compounds (Morales and Munne-Bosch, 2019). Therefore, HPLC, as used in our study, generates a precise, sensitive, and reproducible approach for quantifying lipid peroxidation (Mateos et al., 2005). Consequently, the quantification of lipid peroxidation should be based on its secondary product, MDA, as opposed to the more volatile LOOH concentrations. Furthermore, MDA-modified LDL rather than LOOH-modified LDL is a key factor contributing to degenerative processes (e.g. increasing atherogenicity) (Lankin et al., 2023).

Nevertheless, the strong inverse correlations of the phenome features of SDMD and FE-SDMD with LOOH suggest that lowered LOOH may have some relevance. Moreover, in at least one other study, decreased LOOH was linked to depressive (prenatal) symptoms (Roomruangwong et al., 2017). The primary oxidation products of lipid oxidation are lipid peroxide radicals (LOO[). The former compound may decompose easily into ^1^O2 (the Russell mechanism), cause lipid peroxidation propagation, and is transformed into aldehydes or LOOH (Lankin et al., 2023). Thus, lowered LOOH levels may indicate a more profound conversion of LOO[ into aldehydes, singlet molecular oxygen, and lipid peroxide propagation. The latter may be further activated by increased ONOO^−^ (peroxynitrite) and hypochlorous acid (HOCl) (Lankin et al., 2023) which are both associated with MDD (Maes et al., 2011). Hence, increased MDA coupled with lower LOOH may indicate more detrimental effects of the lipid peroxidation pathway.

### 4.3. O&NS and the clinical features of SDMD

The second major findings of this study are that a) there was a significant and positive correlation between the O&NS biomarkers and psychological stressors, such as ACEs and NLEs; and b) O&NS pathways accounted for a substantial proportion of the variance in clinical rating scale scores for depression severity, brooding, and the phenome, even when ACEs and NLEs were controlled for. Some evidence suggests that psychosocial stressors might contribute to increased levels of O&NS and reduced antioxidant defenses (Moraes et al., 2018). Physical neglect has a significant predictive effect on elevated O&NS indicators, as measured by a composite of LOOH + SOD + NOx, according to the authors of this study. In contrast, sexual abuse was associated with a lowered antioxidant composite score, namely zinc + albumin + and -SH groups. In humans, psychosocial stressors are accompanied by increased O&NS (Maes et al., 2011; Moylan et al., 2014) and brain regions of chronic mild stress (chronic social isolation, and restraint stress) in animal models demonstrate increased lipid and protein oxidation and decreased antioxidant levels (Kubera et al., 2011). Thus, stress-induced O&NS contributes to depression in conjunction with immune and neurotoxic pathways.

Most importantly, our results suggest that the effects of O&NS on brooding, the severity of depression, and the phenome of depression remained statistically significant, even after controlling for ACEs and NLEs. The results of this study suggest that the combination of O&NS toxicity and NLEs can substantially predict the manifestation of SDMD, with a part of the effects of NLEs and ACEs being mediated via O&NS pathways. The present study attributed an estimated 51.7% - 55.1% of the variance in the phenome of SDMD and FE-SDMD to the combined influence of NLEs, ACEs, and the O&NS index, indicating that the phenome of this disorder is largely predicted by a combination of physiological and psychological stressors.

We have previously established that O&NS pathways may partially mediate the effects of ACEs on the MDD phenome (Maes et al., 2018a; 2021). Maes et al. (2018b) discovered in a previous investigation that ACEs can predict the phenome of MDD, with a part of these effects being mediated by compromised antioxidant defenses. Further potential mechanisms by which ACEs exert their effects on the manifestation of MDD include the microbiome, increased atherogenicity, T cell activation, and immune-associated neurotoxicity (Almulla et al., 2023; Maes et al., 2023b; Rachayon et al., 2023).

### 4.4. O&NS and neurotoxicity

As discussed in the previous sections, prior research demonstrated unequivocally that as the severity of staging increases (i.e., treatment resistance, chronicity, and expanding ROI), antioxidant defenses diminish, and O&NS levels rise. Furthermore, antioxidant defenses may

be impacted by O&NS processes, as an increase in ROS/RNS production may result in a corresponding rise in plasma antioxidant consumption (Maes et al., 2008; 2011). A pernicious cycle may ensue in which the organism’s antioxidant systems fail to defend against ROS/RNS and oxidative challenges as antioxidant levels continue to decline (Moylan et al., 2014).

Importantly, the brain is highly sensitive to increased ROS/RNS. Physiological concentrations of ROS and RNS are crucial for numerous physiological processes (Valko et al., 2007). However, when ROS levels surpass the antioxidant capacity of an organism, cellular components may be oxidatively damaged (Aitken and Roman, 2008). ROS/RNS overproduction induces O&NS, specifically in the brain, the site of highest mitochondrial activity (Priyanka et al., 2013).

This modulation can lead to an increase in ROS/RNS production within mitochondria (Di et al., 2009). It has been conclusively established that increased ROS and RNS in the extracellular space contribute to the pathogenesis of MDD and are a primary cause of neurotoxicity processes (Bhatt et al., 2020; Maes et al., 2011; Moylan et al., 2014). Mitochondria may be harmed by elevated concentrations of MDA and other aldehydes, including 3-hydroxynonenal, which suppress levels of mitochondrial GSH and SOD. In addition, MDA has the potential to impede mitochondrial respiratory processes, complexes I, II, and V, as well as the mitochondrial membrane potential, resulting in neurotoxicity and subsequent mitochondrial damage (Long et al., 2006; 2009). These processes are known to be associated with MDD (Maes et al., 2011).

#### Limitations

This study would have been even more interesting if we had measured other O&NS indicants such as the glutathione system and glutathione peroxidase, superoxide dismutase, xanthine oxidase, nitrosylation, etc. It may be argued that the sample size is not large. Nevertheless, the a priori study sample was based on power calculation, and the primary outcome of this study was performed at a power of 1.0.

## Conclusions

Both SDMD and FE-SDMD are characterized by increases in oxidative stress (MDA and NOx) and attenuated antioxidant defenses (lowered TRAP). The combined effects of increased NLEs and ACEs and oxidative and nitrosative stress are strongly associated with the phenome of SDMD and FE-SDMD. This indicates that the effects of psychological stressors combined with O&N stressors predict the phenome of SDMD and FE-SDMD. Given the discussion above (see section 4.4), we may conclude that aldehyde and NO formation are new drug targets to treat FE-SDMD and to prevent any further decline in antioxidant defenses and aggravation of oxidative stress damage and neurotoxicity. In fact, given our negative findings on PON1 activities, it may be hypothesized that lowered PON1 activity because of increased O&NS due to psychosocial stressors is another drug target to prevent an ever-increasing ROI.

## Data Availability

All data produced in the present study are available upon reasonable request to the authors

## Ethical approval and consent to participate

The research project (IRB no.351/63) was approved by the Institutional Review Board of Chulalongkorn University’s institutional ethics board, Bangkok, land, which follows the International Guideline for Human Research protection as required by the Declaration of Helsinki, The Belmont Report, CIOMS Guideline and International Conference on Harmonization in Good Clinical Practice (ICH-GCP). All participants signed the appropriate institutional informed consent forms before data collection.

## Availability of data and materials

MM will reply to reasonable requests for the dataset used in the current study after it has been fully utilized by all authors.

## Credit author’s contributions

FFB and MM wrote the first draft. AV and MM carried out the current study’s design. The data was gathered by AV. Analyses were performed by FFB, LOS, APM, AKM and DSB. Statistical evaluation was performed by MM. All authors contributed to the editing of the work, and they have all given their consent for submission of the completed version.

## Acknowledgments

Not applicable

